# Machine Learning-Enabled Prediction of Speech Perception Improvement Based on Diffusion Tensor Imaging of Young Cochlear Implant Candidates

**DOI:** 10.1101/2022.11.22.22282362

**Authors:** Xiujuan Geng, Patrick CM Wong, Elizabeth Tournis, Maura Ryan, Nancy M Young

## Abstract

Prediction of improvement in speech perception after cochlear implantation (CI) is clinically important to optimize pediatric habilitation. Conventional methods using non-neural measures do not permit accurate prediction on the individual child level. In this study, we investigate whether white matter patterns detected by diffusion tensor imaging (DTI) magnetic resonance imaging (MRI) predict speech perception improvement. Pre-surgical DTI of CI candidates was compared to matched normal-hearing (NH) children to determine cortical regions affected by hearing impairment. Speech Recognition Index in Quiet was measured at baseline and 6 months post implantation to compute improvement in speech perception. Machine learning prediction of speech perception improvement (high or low) was performed using non-imaging and DTI white matter characteristics of whole, affected and unaffected brain. Affected and unaffected white matter regions were determined by comparing DTI multi-voxel pattern similarity maps of white matter integrity indices between CI and NH. Abnormal white matter patterns were found throughout brain of CI candidates. Prediction of 6-month post-CI improvement accuracy, sensitivity and specificity using unaffected regions (0.86, 0.91, 0.80, respectively) and whole brain white matter (0.85, 0.91, 0.80, respectively) yielded similar results, and were more predictive than regions affected by hearing impairment (0.72, 0.74, 0.70, respectively) or non-imaging features (0.67, 0.55, 0.78, respectively). Findings support that presurgical neural white matter pathways, especially in the association auditory and cognitive regions not affected by auditory deprivation, play a critical role in speech development after CI and are more predictive of outcome than traditional non-neural variables such as age at implant.

## Introduction

Cochlear implantation, despite its effectiveness, is associated with significant outcome variability. Even when implanted at a young age, only a minority of children achieve language equivalent to normal-hearing children (Niparko et al. 2010). Studies of implanted children show group language performance at about the 15^th^ percentile of normal-hearing children, close to the range of developmental language disorder (Nittrouer et al. 2016). Several decades of research show that patient characteristics, in particular age at implant and residual hearing, impact language outcome. However, prediction based on these characteristics is not accurate on the individual level, only for groups when sample size is large.

In 2017, Wong et al proposed an approach to language learning inspired by personalized medicine in which genotypic information is used to develop effective individualized drug regimens (Wong et al. 2017). When applied to children who are cochlear implant (CI) candidates, prediction would enable identification of children at risk to be low performers, providing the opportunity to optimize type and intensity of behavioral therapy.

This approach has potential to improve outcome and cost effectiveness by efficiently allocating therapy resources. Our research uses brain anatomy from pre-CI magnetic resonance imaging (MRI) to improve prediction of language outcome. We hypothesize that brain anatomy provides neural predictors superior to traditional patient characteristics such as age at implant and residual hearing. The rationale for superiority of neural predictors is that central processing is essential to outcome given: 1) The sparse information provided by CI in comparison to normal acoustic hearing, creating a burden on central auditory processing; and 2) the potential impact of auditory deprivation on brain development (Kral and Sharma 2012, Kral 2013, Sharma et al. 2002).

Our group has previously demonstrated that neuroanatomical brain measures taken from standard pre-surgical T1-weighted MRI, when combined with machine learning analytics, can predict CI speech perception at 6-months with greater accuracy than traditional patient characteristics (Feng et al. 2018). That longitudinal study also evaluated competing hypotheses as to whether brain regions impacted or unimpacted by auditory deprivation would contribute most to improvement. The results support the hypothesis that preservation of brain areas unaffected by auditory deprivation, especially higher-level auditory processing areas, contributes to enhanced outcome (Feng et al. 2018).

The current study employs diffusion tensor imaging (DTI). DTI is an MR technique that takes advantage of restricted free diffusion properties of water molecules along axons. It is highly sensitive to microstructural changes not identifiable by standard MRI and is useful in understanding fiber tract connections between brain regions. The study goal is to determine if DTI combined with machine learning predictive models can forecast speech perception before and after CI.

## Materials and Methods

### Subjects

The study was Institutional Review Board approved (IRB 2011-1464 and 2018-1959) and parental consent was obtained for 52 (32 female) CI candidates between 2011 – 2020.

Fourteen children used bilateral CIs. Comparison group was 31 (17 female) age- and sex-matched normal hearing (NH) children from an NIH shared repository (PedsMRI; https://pediatricmri.nih.gov/nihpd/info/index.html). HL group mean age at MRI=16.1±8.6 months; NH group=16. 8±8.8 months. Age at implant=19.3±8.7 months. HL group received standard MRI with additional DTI. Exclusion criteria included: developmental delay and conditions expected to impact outcomes; gross brain malformation, severely malformed cochlea and cochlear nerve deficiency. Children in the NH group were matched by age at scanning with HL group (p=0.867) and sex (p=0.55).

### Auditory and Speech Measures

The primary outcome measure was the Speech Recognition Index in Quiet (SRI-Q) used in the Childhood Development after Cochlear Implantation Study (Niparko et al. 2010, Fink et al. 2007). The SRI-Q includes the Meaningful Auditory Integration Scale (MAIS) and Infant Toddler MAIS (IT-MAIS), a parental report and criterion-referenced scale of children’s auditory skills that is not ear-specific. The IT-MAIS/MAIS was completed before CI and at 6-months after activation. Scores 6-month post-activation were available in 32 (female=19). Aided speech awareness threshold (SAT) was used as a proxy for residual hearing.

### MRI Acquisition and Processing

Scans for HL group were acquired on a 3T Siemens scanner (MAGNETOM Skyra or Vida) using an MPRAGE sequence. Scanning parameters were optimized to improve signal-to-noise ratio. Scanning parameters were as follows: for T1w: TE = 2.60–3.25 ms, TR = 1,900 ms, flip angle = 9°, matrix = 256 × 256, 176 slices of 0.9-mm thickness, voxel size = 0.9 × 0.9 × 0.9 mm; for DTI: TE=84 ms, TR=8s, voxel size=2 × 2 × 2mm^3^; b=1000s/mm^2^; number of diffusion gradient directions=64. The T1-weighted and DTI scans of the NH group were downloaded from the NIH sharable database. Parameters from the NIH database for the normal-hearing children included: for T1w: TE = 10 ms, TR = 500 ms, flip angle = 90°, matrix = 256 × 192, 30–60 slices of 3-mm thickness, voxel size = 1 × 1 × 3 mm; for DTI: voxel size=2.5 × 2.5 × 2.5 mm^3^; number of gradient directions=110 or 120.

Images were processed using FMRIB Software Library (FSL) tools (Jenkinson et al. 2012), SPM 12 (Wellcome Department of Imaging Neuroscience, London, United Kingdom; www.fil.ion.ucl.ac.uk/spm/), CoSMoMVPA toolbox (Oosterhof et al. 2016), and in-house scripts. Images from HL and NH groups were visually inspected to ensure whole image coverage. Eddy current correction was first performed. Tensor images were then estimated and maps of fractional anisotropy (FA), and radial (RD) and axial diffusivities (AD) were computed afterwards. Due to the absence of DTI atlas for infant brains, we affine aligned JHU T2 template in FSL to the UNC 1-year infant T2 brain atlas (Shi et al. 2011) and applied this transformation matrix to obtain the JHU FA template in the 1-year-old brain space (JHU_1yr). Next, the FA maps of individuals from both groups were affine aligned and non-linear warped to this JHU_1yr template using FSL. Corresponding transformation matrix (for affine alignment) and displacement fields (for non-linear registration) were applied to the RD and AD maps to warp them into the same template space.

### Multi-Voxel Pattern Similarity (MVPS) Analysis

MVPS analysis considers neighboring distributions and provides stronger detection power compared to uni-voxel analysis (Kriegeskorte et al. 2006). Different acquisition protocols from two population pools may affect direct comparison of the uni-voxel based DTI properties between the two groups. Therefore, we conducted MVPS analysis of FA, RD and AD maps on white matter across whole brain. The average of the registered FA maps were computed and the regions with FA values greater than 0.15 were set as white matter. The computation of MVPS of FA, RD and AD maps was done using a searchlight approach with a three-voxel-radius sphere centered on each voxel across white matter to define the local spatial extent. The local white matter pattern similarity was calculated for each of local spheres using Pearson correlation of each pair of individuals between HL and NH groups and within the NH group. For each child in HL group, there were 31 between-group similarity maps (from this child in HL group to all 31 children in NH group). These maps were averaged to produce each child’s between-group similarity map. Similarly, the between-group similarity map for a child in NH group was averaged across 52 between-group similarity maps. The within-group similarity map for an individual in NH group was the average of the similarity maps from this child to all other children in the NH group.

### Machine-learning Classification

Two sets of classification for predicting speech perception using SRI-Q were conducted at baseline and 6 months after CI activation. Median split was used to separate children into high and low speech performance groups at baseline. Similarly, the improvement of speech outcomes (6-months post-CI – baseline scores) was separated into high and low improvement groups using median split of the score differences. For each set of predictions, three prediction models were used: the input features including all MVPS values from 1) whole brain white matter; 2) only affected regions; and 3) only regions without significant abnormalities of the between-group and within-group MVPS maps. The MVPS maps of FA, RD and AD from each child were converted into a matrix consisting of S rows and V columns, where S is the number of subjects and V is three times of the number of voxels from regions used in the prediction model, with each voxel containing MVPS values of FA, RD and AD. S = 52 for prediction of baseline speech performance; and S=32 for prediction of 6-month speech improvement.

Machine learning classification was performed using the support vector machine (SVM) from Scikit-learn (Pedregosa et al. 2011). To avoid overfitting using a large number of predictive features, leave-one-fold-out cross validation procedure was employed. At the inner level, we iteratively chose best features using SelectKBest together with the kernel type (‘linear’ and ‘rbf’ for linear and nonlinear kernels respectively), and other two important parameters C and Epsilon to select the best “K” features and the best parameters using the training folds, which were sent to the one-fold for testing. Once the model was chosen, this cross validation using the selected features and parameters was repeated 10,000 times to generate the distribution of the classification performance. Classification accuracy (ACC), sensitivity and specificity, and area under the curve of the receiver operating characteristic curve (AUC) were calculated and statistically compared under various conditions (whole white matter vs. affected vs. unaffected brain regions). The affected templates were built by combining the voxels that showed significant group difference from MVPS analyses using FA, RD and AD maps. The unaffected templates were built by including the voxel that did not show significant group difference from the MVPS analyses.

### Statistical Analysis

To analyze white matter reorganization for HL group, statistical tests between 52 between-group MVPS maps (for HL group) and 31 within-group MVPS maps (for NH group), controlling for age and sex, were performed. The within-group MVPS maps from NH group were considered as a normal baseline. Two-sample t-tests were used to assess significant differences between and within-group for each voxel. After statistical correction for multiple voxels, significant group difference maps were thresholded with the corrected p < 0.05 using a standard nonparametric permutation method used in neuroimaging studies to perform multiple comparison corrections.

Within HL group, comparison of pre- and post-CI SRI-Q scores was performed using one-way ANOVA with repeated measures, with age and sex as covariates. Spearman rank correlations were computed between all of the nonneural measures. The ANOVA and correlation analyses were conducted using SPSS (version 22). These tests were two-tailed, with an a priori assumption of significance at p ≤ 0.05 but corrected for multiple tests when appropriate.

To determine whether classification accuracy of each predictive model was significantly better than chance, a nonparametric permutation procedure was used to generate a chance distribution (null hypothesis). To achieve this, the group labels were randomly assigned into two new groups and the classification procedure was repeated. This randomization procedure was repeated 10,000 times, and the 95^th^ percentile points of each distribution were used as the critical values for a one-tailed t test of the null hypothesis with p=0.05. To compare the performance of different predictive models, data were randomly split with leave-one-fold-out with one-fold for testing once the best features and parameters were chosen, and repeated the procedure 10,000 times to generate a performance distribution for each predictive mode. Accuracy, area under the curve, sensitivity and specificity of each predictive model were compared against the null distribution. To compare the performance between different predictive models, e.g., comparing models using non-neural vs. imaging features, we also used 95^th^ percentile points of the model distribution as the critical values to calculate statistics.

## Results

### White matter reorganization related to auditory deprivation

Changes in white matter integrity, including myelination and reorganization properties, can be inferred from the MVPS maps by comparing MVPS differences between HL and NH groups. Brain regions where there are significant differences between children in HL and NH groups are illustrated and named in Fig. 1. Many of these regions include primary and associated auditory cortex and other higher cognitive cortices.

**Fig. 1.**
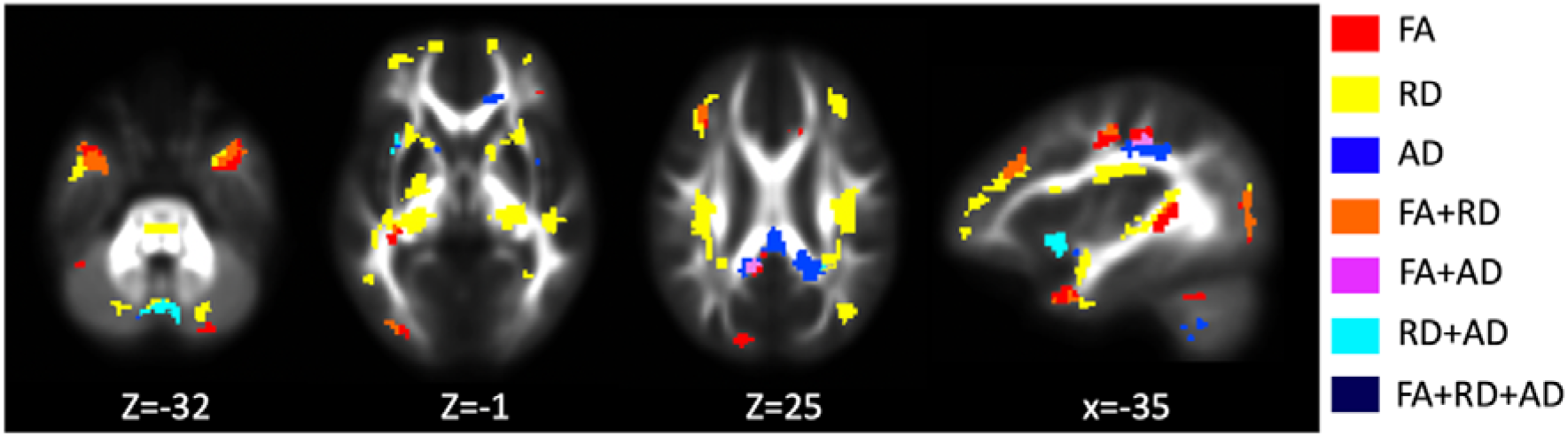
Group differences (hearing loss and normal hearing) in MVPS analysis of white matter maps FA= fractional anisotropy; RD=radial diffusivity; AD= axial diffusivity. Significant differences in the MVPS maps are shown in colored regions. Regions with major differences between the HL and NH groups include: brain stem on cortical spinal tract (CST); bilateral pyramid of vermis of cerebellum; bilateral superior and middle temporal pole region on uncinate fasciculus (UF), and inferior longitudinal fasciculus (ILF); bilateral posterior superior temporal gyrus (pSTG) on ILF; bilateral prefrontal regions on forceps minor; bilateral dorsal lateral frontal regions, and globus pallidus on anterior thalamic radiation (ATR), CST and superior longitudinal fasciculus (SLF); bilateral postcentral gyrus on SLF; bilateral posterior cingulate gyrus (PCG) on cingulum; left middle occipital gyrus on ILF, inferior fronto-occipital fasciculus (IFOF) and forceps major; and right occipital region on IFOF.

### Predicting baseline and 6 month speech perception performance

Statistical analysis, controlling for age and sex, showed significant improvement in SRI-Q at 6 months post-activation compared with pre-CI scores [Mean pre and post=17.9 and 81.0, respectively; F(1,62)=63.54, p<0.001]. Improvement was calculated using the paired differences between post- and pre-CI scores (Fig. 2). There was no significant correlation between age at CI and the pre-CI SRI-Q (Rho=-0.125, p=0.496) or post-CI scores (Rho=-0.041, p=0.823). Residual hearing was significantly correlated with socioeconomic status (Rho=-0.467, p=0.011), whether implantation was unilateral or bilateral (Rho=0.483, p=0.008), and baseline MAIS/IT-MAIS scores (Rho=-0.627, p<0.001). The 6-month post-CI scores were correlated with baseline scores (Rho=0.377, p=0.034), and the 6-month improvement scores (Rho=0.782, p<0.001), but there was no significant correlation between baseline and 6-month improvement scores.

**Fig. 2.**
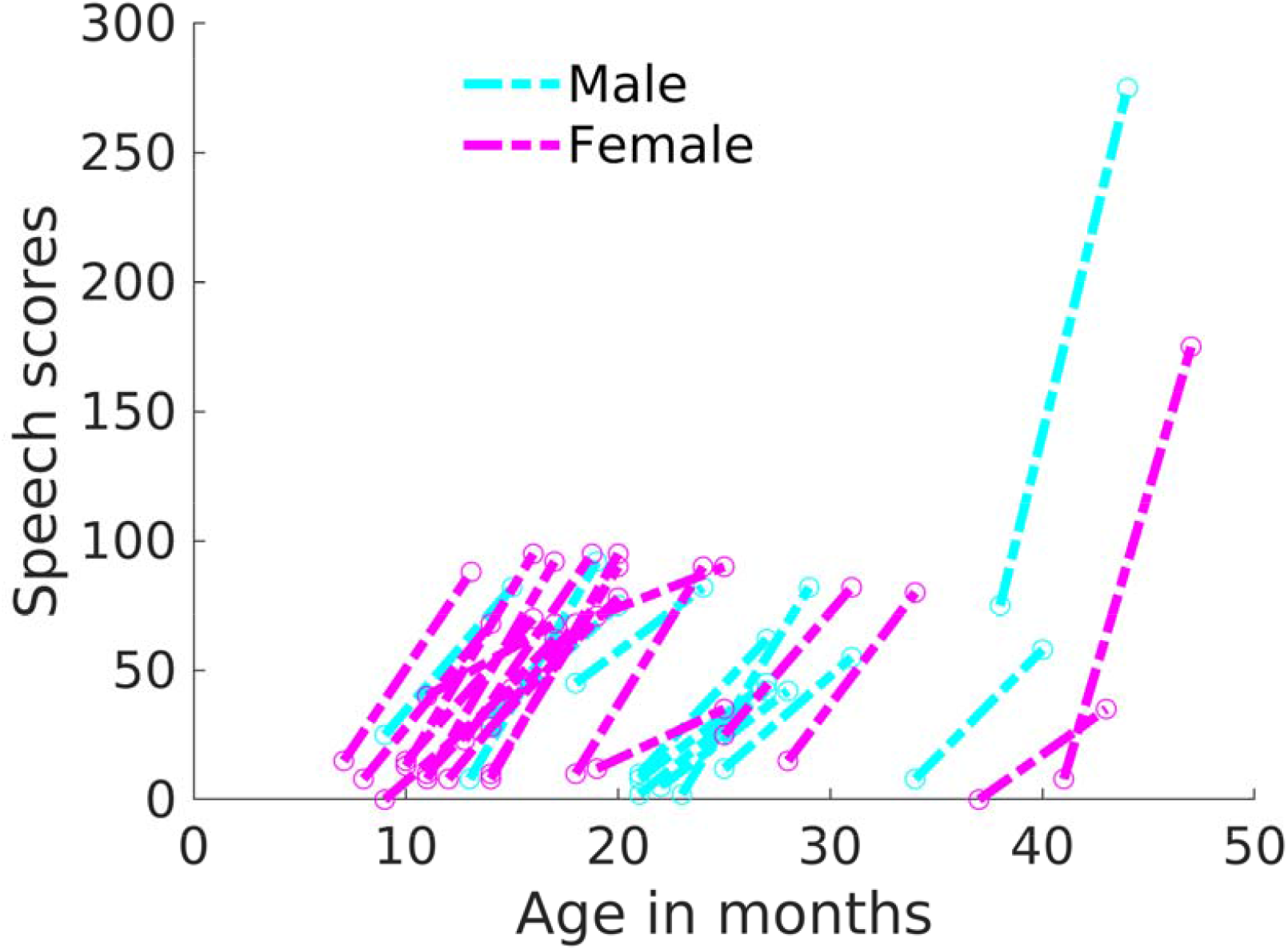
Scatterplot of longitudinal changes (baseline to 6-months) in speech perception score (SRI-Q) for 32 implanted children The two scores (baseline and 6 months after cochlear implant activation) are linked by a dashed line for each subject and shown separately for males and females.

Imaging measures, including the MVPS of FA, RD and AD maps, were the input features to the machine learning for prediction of baseline scores and 6-month improvement. Classification was also performed using age, sex and residual hearing as predictors. The accuracy of the models using whole, affected, and unaffected brain as well as the non-imaging factors in classification of improvement are shown in Fig. 3. Although non-imaging features can generate classification accuracy significantly higher than the null distribution (ACC=0.676, p=0.0025 for baseline, and mean ACC=0.672, p=0.007 for 6-month improvement), performance was not as good at prediction as neural imaging measures, with ACC using non-imaging features significantly smaller than using neural predictors (p=0.05 for baseline, p<0.001 for 6-month improvement). Accuracy, sensitivity, specificity, and AUC under various classification conditions are provided in Table 1.

**Fig. 3.**
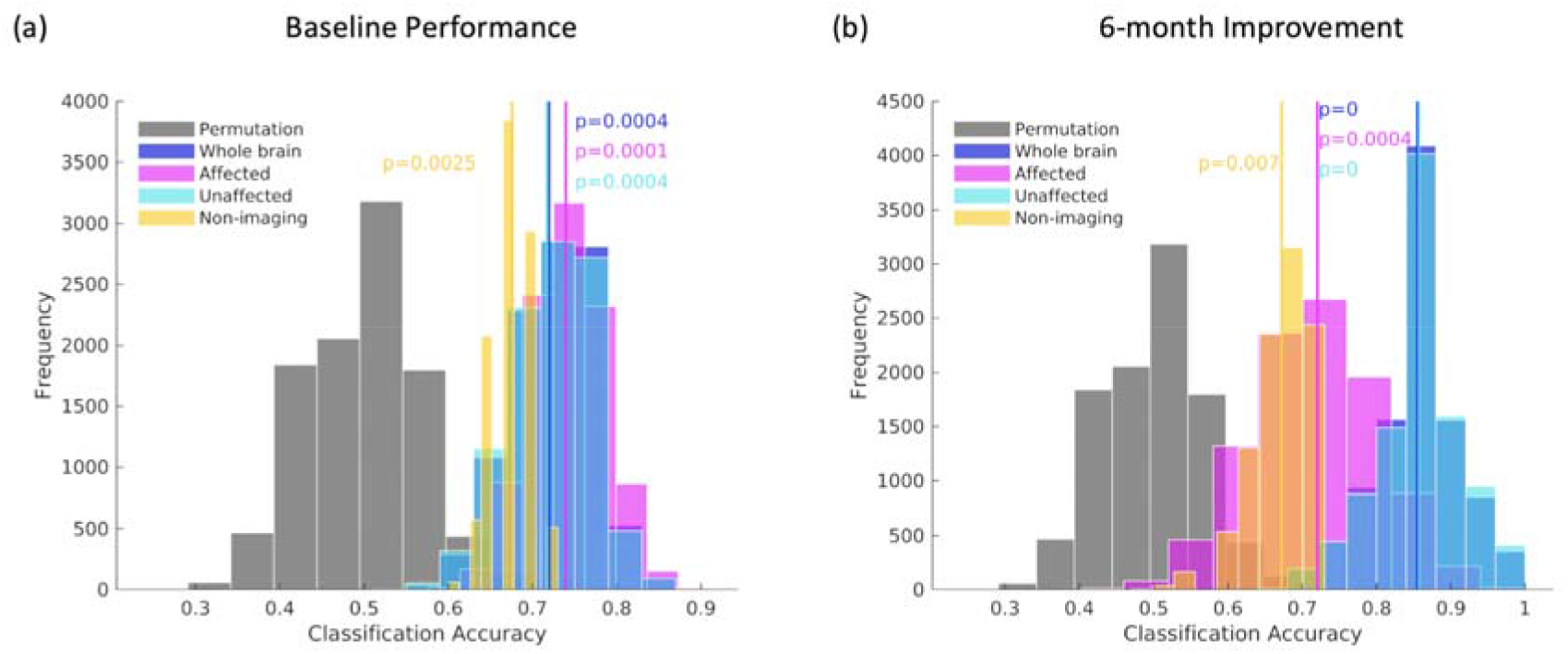
Classification of baseline (a) and 6-month improvement (b) performance using non-imaging and imaging features Classification of baseline (a) and 6-month improvement (b) performance using non-imaging features (including age, sex, SES, and residual hearing measured by SAT), whole brain MVPS maps, MVPS maps on affected brain regions, and MVPS maps on unaffected brain regions. The classification procedure was repeated 10,000 times for estimating the distribution of classification accuracy. The classification accuracies of all models are significantly better compared to the null distribution.

**Table 1.** Mean (SD) speech perception prediction performance of white matter pattern similarity at baseline and 6-month using various models.

|  | Whole Brain | Affected | Unaffected | Non-imaging |
| --- | --- | --- | --- | --- |
| **Baseline** |  |  |  |  |
| ACC | 0.72 (0.05) | 0.74 (0.05) | 0.72 (0.05) | 0.68 (0.02) |
| AUC | 0.73 (0.05) | 0.78 (0.04) | 0.73 (0.05) | 0.78 (0.01) |
| Sensitivity | 0.75 (0.08) | 0.78 (0.07) | 0.70 (0.08) | 0.64 (0.03) |
| Specificity | 0.69 (0.08) | 0.70 (0.08) | 0.73 (0.08) | 0.71 (0.03) |
| **6-Month** |  |  |  |  |
| ACC | 0.85 (0.06) | 0.72 (0.09) | 0.86 (0.06) | 0.67 (0.04) |
| AUC | 0.94 (0.04) | 0.87 (0.07) | 0.94 (0.04) | 0.55 (0.03) |
| Sensitivity | 0.91 (0.06) | 0.74 (0.10) | 0.91 (0.06) | 0.55 (0.04) |
| Specificity | 0.80 (0.09) | 0.70 (0.10) | 0.80 (0.09) | 0.78 (0.08) |

SD: standard deviation; ACC: accuracy; AUC: area under curve.

### Overlapping regions of altered brain and predicting baseline and 6-month improvement

Conjunction maps were generated between the affected regions and the predictive regions for baseline performance and 6-month improvement performance (Fig. 4). Some of the baseline predictive regions are overlapped with the white matter abnormalities while some are not.

**Fig. 4.**
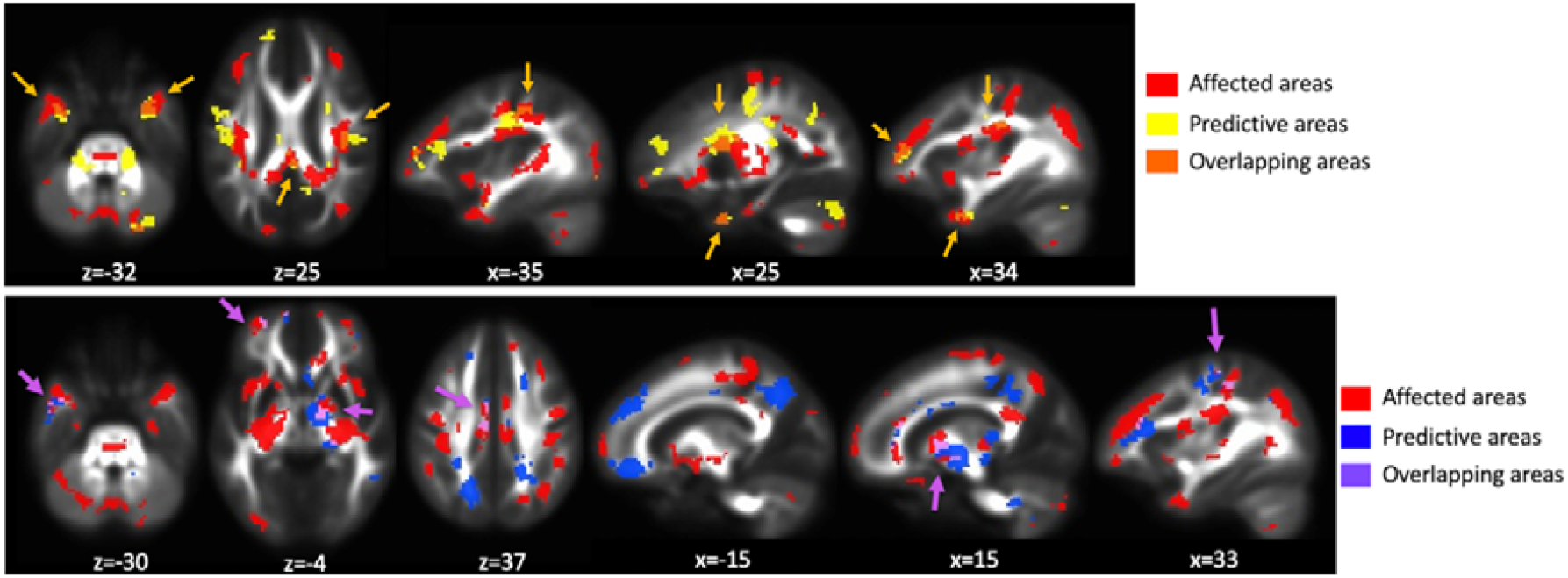
Regions predicting baseline (top) and 6-month improvement (bottom) speech perception performance overlap with hearing loss white matter abnormalities compared with normal hearing The arrows indicate overlapping areas between the predictive and abnormal (compared to normal hearing) regions.

The overlapping regions include: bilateral superior/middle temporal gyrus/uncus on inferior longitudinal fasciculus (ILF); bilateral postcentral gyrus on superior longitudinal fasciculus (SLF); right middle frontal gyrus on right anterior thalamic radiation (ATR), right posterior cingulate and uncus on cingulum; right putamen/caudate/lentiform nucleus on cortical spinal tract (CST). Some of the baseline predictive regions did not overlap with the group difference including: bilateral brain stem; left precentral gyrus on SLF, left prefrontal gyrus on forceps minor, left inferior frontal gyrus on ATR, right middle frontal gyrus on inferior fronto-occipital fasciculus (IFOF), and right cingulate/middle frontal gyrus on CST/SLF.

The predictive regions of 6-month improvement are also partially overlapped with the abnormal white matter regions, but most of the predictive regions are from unaffected areas. The overlapping regions are: left middle temporal gyrus on ILF; left superior frontal gyrus on ILOF; left cingulate gyrus on cingulum; right globus pallidus on ATR; and right postcentral gyrus on SLF.

We have summarized the affected and predictive regions by calculating the overlapping voxels of these regions with 20 major white matter tracts provided by FSL. The affected white matter spread across most of the major tracts (Fig. 5), and the left hemisphere showed more affected area voxels than the right hemisphere for most of the tracts. The most predictive regions of baseline were primarily those along left SLF, right CST, right SLF, and right ATR. Regions most predictive of 6-month improvement were primarily located along right ATR, IFOF and SLF, forceps minor and left cingulum.

**Fig. 5.**
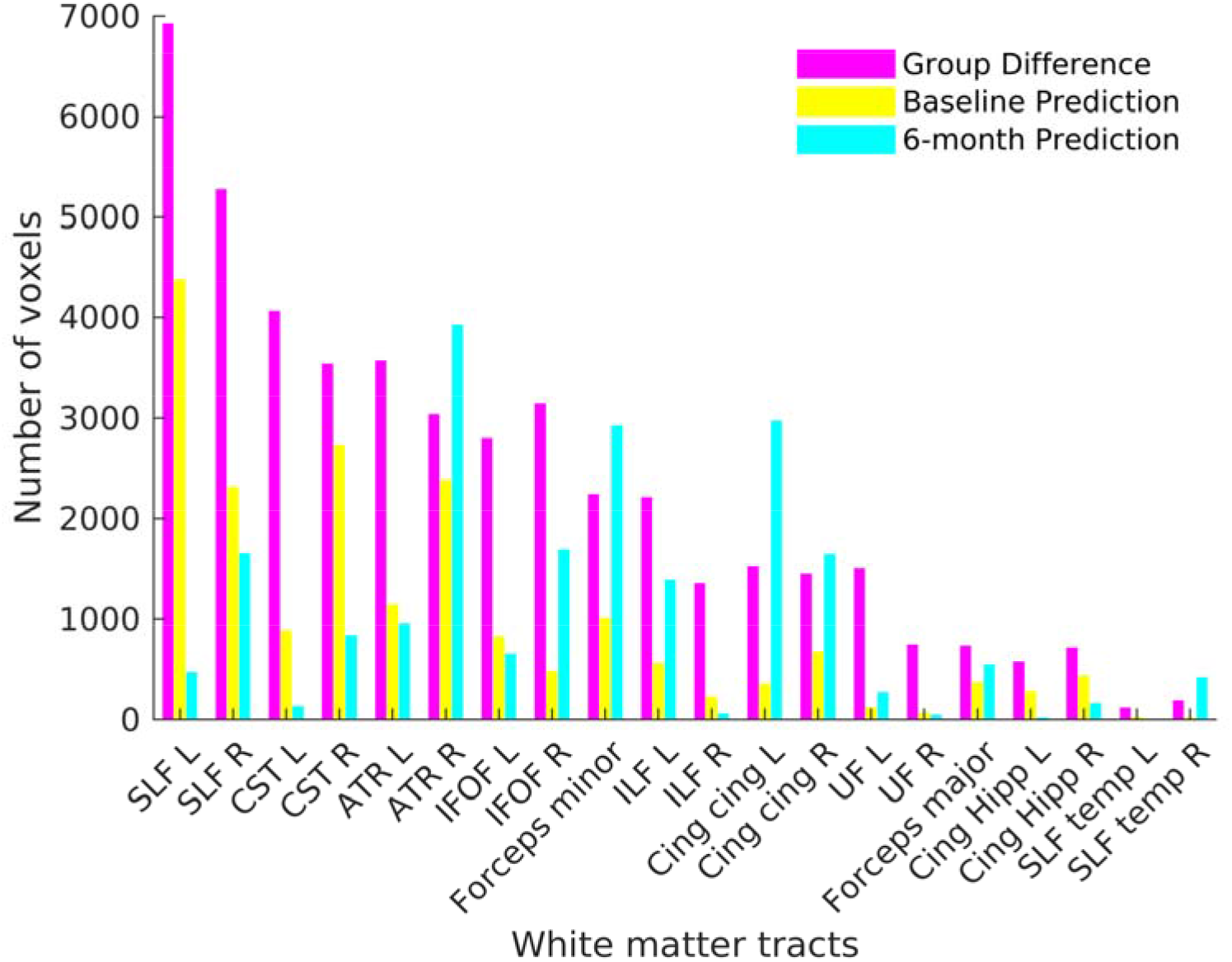
Bar plot of cluster sizes (for group difference, predictive for baseline and 6-month post-CI speech improvement) overlapped with the 20 white matter tracts provided by FSL SLF=superior longitudinal fasciculus, CST=cortico-spinal tract, ATR=anterior thalamic radiation, IFOF=inferior fronto-occipital fasciculus, ILF=inferior longitudinal fasciculus, Cing cing=cingulum in the cingulate gyrus, UF=uncinate fasciculus, Cing Hipp=cingulum in the hippocampal formation, SLF=superior longitudinal fasciculus. R=Right, L=Left.

## Discussion

Cochlear implantation is the most effective treatment available for children with significant bilateral sensorineural hearing loss not adequately ameliorated by amplification (Kral and O’Donoghue 2010). However, speech and language outcomes after CI are variable and hard to predict (Niparko et al. 2010). Prediction of language outcome on the individual child level could lead to customized treatment to improve the language trajectory of those at risk to develop far below their normal-hearing peers (Wong et al. 2017).

Studies using traditional non-neural variables to predict outcomes in adult CI recipients have demonstrated that these variables explain only 10-20% of variation in speech perception outcomes (Zhao et al. 2020, Lazard et al. 2012). Few studies have shown success predicting which children with HL will develop age-appropriate language. Several studies using fMRI to predict language employed auditory task-based fMRI (Deshpande et al. 2016, 2018, Tan et al. 2015). However, use of auditory tasks for children with significant HL introduces confounding factors. Similarly, other fMRI studies introduced a confounding variable by using sedation known to interfere with cerebral metabolism (Peltier et al. 2005, Hamilton et al. 2017).

The limitations of fMRI studies highlight the need for another approach. DTI is another means to detect neuroanatomic differences that may be useful in forecasting language at the individual level (Floel et al. 2009). It has been used to compare white matter structure of deaf and hearing individuals (Simon et al. 2020, Ratnanather 2020). Two studies used pre-surgical DTI to predict CI outcomes (Huang et al. 2015, Wang et al. 2019), but children in these studies, done in Mainland China, had a much older age at implant (3 to 4 years) than in the US. Nonetheless, they provided important proof-of-concept data suggesting DTI could be useful for prediction.

The present study investigated whether pre-surgical DTI neural patterns coupled with machine learning analytics can predict differences in speech perception improvement, and if so, how that compares to use of traditional non-neural factors such as age at implant and residual hearing. Residual hearing was significantly correlated with baseline speech perception scores, as would be expected, but was not correlated with 6-month scores or improvement classification. No significant correlations between age at implantation and baseline speech perception or 6-month improvement were found, perhaps due to the relatively narrow age range. Machine learning prediction using non-imaging characteristics did predict baseline speech perception and 6-month improvement significantly better than a null model. However, the prediction accuracy of machine learning using DTI outperformed traditional characteristics for prediction of baseline and speech perception improvement. This finding is consistent with our previous study demonstrating that neural measures from T1-weighted MRI scans predicted speech perception improvement with greater accuracy than traditional patient characteristics (Feng et al. 2018). The finding that brain regions unaffected by auditory deprivation are most predictive of post-CI improvement is also consistent with our previous study, providing converging evidence of the importance of preservation of higher cognitive regions.

The brain’s neural sensory system develops before birth (Graven and Browne 2008). When auditory sensory input is deficient, neural reorganization may occur in infants and children (Bavelier and Hirshorn 2010, Hribar et al. 2020). White matter abnormalities caused by HL have been found in young children in studies using whole brain white matter analysis (Park et al. 2018, Jiang et al. 2019). Compared to NH children, children with HL age 1 to 4 years have a smaller number of white matter tracts. The current study found many of the same abnormalities. Our findings, together with other studies, indicate that congenital HL causes white matter deficits not only in primary auditory pathways, but also in the secondary auditory and higher-level cognitive pathways. The impact of auditory deprivation is not limited to impaired ability to perceive and process auditory input, but also may alter processes such as auditory-related emotional response, creation and recall of memories, learning, production and comprehension of language (McAdams and Bigand 1993). DTI studies using microstructural measures such as FA consistently observe altered microstructure in auditory areas not typically found in T1-weighted MR studies. This difference can be attributed to the better sensitivity of DTI to detect subtle structural changes that precede volumetric changes evident on T1-weighted MRI (Hribar et al. 2020).

Instead of the primary auditory regions, white matter pathways such as the superior longitudinal fasciculus, anterior thalamic radiation and cingulum, underlying association and higher-order cognitive brain regions are more likely to predict individual speech development after CI in young children. By making conjunction maps between the predictive and group difference maps, we found that the majority of predictive areas for both baseline and post-CI improvement are from unaffected brain regions. This finding provides further evidence that preservation of brain areas unaffected by auditory deprivation influences post-CI speech perception.

A number of studies have been done using presurgical neural measures such as glucose metabolism in the auditory and related cortices with PET to predict post-CI outcome (Lee et al. 2001, Giraud and Lee 2007). However, they included implanted children and adults across a large age range. Children late implanted (after age 7 years) have less neural plasticity compared to those with early implantation (before age 4 years), and there is a large difference in speech perception of early versus late implanted children (Sharma et al. 2002). Therefore, including older late implanted children may cause overestimation of the effect of brain measures (Lee et al. 2001, Giraud and Lee 2007). Several DTI studies to forecast CI outcomes in children have demonstrated limited prediction (Huang et al. 2015, Wang et al. 2019). However, these studies relied upon narrow pre-selected brain regions, rather than using MVPS to evaluate the cortex. Studies using very limited brain regions may lack power for detecting relationships between imaging and behavioral outcome.

The current study, which unlike prior studies used the advanced technique of MVPS, yielded significant findings suggesting: 1. a greater range of white matter abnormalities in children with severe to profound hearing loss, and 2. unaffected white matter regions are more predictive of individual children experiencing greater improvement in speech perception. One caveat is that due to young age, direct measure of speech perception was often not possible in our subjects. For this reason, the IT-MAIS/MAIS was the only measure consistently available across subjects. This measure, an auditory skills questionnaire, is the lowest level of the SRI-Q hierarchy of auditory skills battery. Future studies of our prediction models with longer-term follow-up are needed including direct measures of speech perception and spoken language.

To summarize, improved prediction of speech perception on the individual child level may be achieved with neural measures from DTI, in comparison to traditional characteristics such as age at implant, when used in conjunction with machine learning analytics.

Preservation of brain unaffected by HL, especially areas of higher-level cortical function, is correlated with more accurate prediction. These findings provide further evidence that brain regions not impacted by HL may significantly contribute to individual difference in speech perception after CI. Knowledge gained from understanding the impact of HL on neural networks and brain microanatomy combined with outcome prediction to identify children at risk not to develop age-appropriate language may permit early behavioral interventions to improve auditory and language outcome.

## Data Availability

All data produced in the present work are contained in the manuscript.

## Acknowledgment

The authors thank Karen I. Berliner, Ph.D. for her considerable help in manuscript editing. The study was supported by the Research Grants Council of Hong Kong (no. 14605119) and NIH grant (no. R21DC016069).

